# COVID-19 Vaccine Perceptions in New York State’s Intellectual and Developmental Disabilities Community

**DOI:** 10.1101/2021.03.19.21253425

**Authors:** Suzannah Iadarola, Joanne F. Siegel, Qi Gao, Kathleen McGrath, Karen A. Bonuck

**Affiliations:** Associate Professor of Pediatrics, Director, Strong Center for Developmental Disabilities, University of Rochester Medical Center, 601 Elmwood Ave, Box 671, Rochester NY 14642; Principal Associate, Department of Pediatrics, Co-Director University Center of Excellence in Developmental Disabilities, Einstein College of Medicine-Montefiore Medical Center, 1300 Morris Park Ave., Bronx NY 10461; Instructor, Department of Epidemiology and Population Health, Einstein College of Medicine-Montefiore Medical Center, 1300 Morris Park Ave., Bronx NY 10461; Candidate, Hunter College, Senior Research Coordinator, Einstein College of Medicine-Montefiore Medical Center, 1300 Morris Park Ave., Bronx NY 10461; Professor, Department of Family and Social Medicine/Department of Pediatrics, Co-Director, University Center of Excellence in Developmental Disabilities, Einstein College of Medicine-Montefiore Medical Center, 1300 Morris Park Ave., Bronx NY 10461

**Keywords:** Vaccines, intellectual disability, developmental disability, COVID-19

## Abstract

**Background:** People with intellectual and developmental disabilities (IDD) are at disproportionate risk for severe COVID-19 outcomes, particularly those living in congregate care settings. Yet, there is limited data on vaccine perceptions in the disability community.

**Objective:** To explore COVID-19 vaccine perceptions in individuals with IDD, their family members, and those who work with them, to inform a statewide vaccine information and messaging project.

**Methods:** A national survey, adapted for the IDD community, was distributed to a convenience sample of IDD organizations throughout New York State, in five languages. Constructs included vaccine intention, reasons for vaccine hesitancy, and trusted sources of vaccine information. Zip code data were used to map respondent location and vaccine preferences.

**Results:** Of n= 825 respondents, approximately 75% intended to or had received the vaccine, across roles (i.e., people with disabilities, family members, direct care workers) and racial/ethnic groups. Greater vaccine hesitancy was reported in younger individuals and those making decisions on behalf of a person with IDD. Concerns included side effects and the swiftness of vaccine development. Black and Hispanic participants had heightened concerns about being an “experiment” for the vaccine. Trusted sources of information included healthcare providers and family members. Respondents who intended/got the vaccine were distributed throughout the state.

**Conclusions:** Vaccine preferences in this New York State disability community sample align with national data. Identified concerns suggest the need for community education that addresses misperceptions. Age and race differences in perspectives highlight the need for tailored education, delivered by trusted messengers.

---

As of Valentine’s Day 2021, there were nearly 28 million cases of COVID-19 in the United States, resulting in nearly half a million deaths. New York State (NYS), an early epicenter of the disease accounts for 5% of the nation’s cases and 10% of COVID-19 death. Despite ubiquitous “we’re all in this together” messaging in the early months, persistent racial, ethnic, and economic disparities have translated to higher infection rates and worse outcomes among persons of color--beyond risk factors associated with age and health comorbidities ^1,2^.

Less well studied are disparities for persons with Intellectual and/or Developmental Disabilities (IDD). The term “IDD” refers to disorders that: begin in childhood; impact an individual’s trajectory of physical, social and/or emotional development, and; may affect multiple body parts of system.^3^ IDD spans a range of conditions such as autism spectrum disorder (ASD), cerebral palsy, Down syndrome, hearing loss, and attention deficit hyperactivity disorder. An estimated 2%-3% of people in the US have some type of IDD ^4^.

Persons with IDD experience a high prevalence of comorbid risk factors (e.g., hypertension, heart disease, respiratory disease) that place them at heightened risk ^5^. Compared to non-disabled persons, those with IDD and related conditions (e.g., Down syndrome and other chromosomal conditions) had the third highest risk of COVID-19 death ^6^. And, intellectual disability was the strongest risk factor for Covid based on data from nearly 6.5 million patients across 547 health care organizations in the US ^7^. COVID-19 risk for persons with IDD is exacerbated for those residing in congregate care settings, given challenges to physical distancing and close proximity to staff ^8^. This has significant implications for the estimated 1-in-5 adults with IDD who live in congregate care ^4^.

In NYS, persons with IDD living in congregate care settings experience more severe COVID-19 outcomes, along with higher case-fatality, and mortality rates ^9^, in a similar pattern to that throughout the United States ^10^. Staff working in NYS congregate care settings became eligible in Phase 1a, while persons with IDD including Down syndrome became eligible in Phase 1a and 1b ^11^. However, across the people with IDD have not been similarly prioritized ^12^. As vaccine access expands to general and IDD populations, vaccine hesitancy may be a barrier to population-level immunity. National data suggests that approximately 25-30% of adults are hesitant to receive the COVID-19 vaccine, even if it were available for free ^13, 14^ Congregate care staff are especially hesitant; one state-wide survey indicated that only 45% of nursing home staff were willing to take an approved COVID-19 vaccine when it became available ^15^.

Promoting vaccine uptake begins with understanding the reasons for vaccine hesitancy. For COVID-19 such reasons include: concerns about side effects, lack of trust given how quickly it was developed, doubts about the vaccine’s efficacy, and beliefs that COVID-19 symptoms are primarily mild ^13, 16, 17^. Historical mistrust in and misconceptions of vaccines within communities of color has been observed, with less intent to get vaccinated in Black survey respondents ^18^. This may be related to Black and Hispanic Americans lacking confidence that the vaccines were adequately tested in their racial and ethnic groups ^19^. Such concerns understandably stem from systematic mistreatment of people of color, but can perpetuate disproportionately severe COVID-19 outcomes within African American/ Black and Hispanic/ Latino communities. Vaccine hesitancy in these communities intersects with the fact that a high percentage of congregate care workers are people of color ^20^. A similar ‘intersectionality’ is seen among parents of color who have children with autism, as they express more vaccine hesitancy (unrelated to Covid-19), compared to white parents ^21^.

Community-focused interventions can increase vaccination uptake ^22^, particularly when: informed by community-specific concerns; targeted toward those who are or are at risk of vaccine hesitancy, and culturally responsive. Additional considerations for people with IDD include ensuring adequate communication around health decisions, inclusion in conversations with providers and through written information ^23^. Preliminary data suggest that, with the authorization of multiple vaccines, there is increased vaccine confidence in communities that have historic hesitancy ^17^, thus outreach and culturally sensitive education around the COVID-19 vaccine is needed in the high risk IDD population.

There is limited data on COVID-19 vaccine perceptions in the IDD community. Though a recent study reports on trust in health information among persons with disabilities ^24^, to our knowledge, no data exist on the IDD community’s COVID-19 vaccine preferences and reasons for them. To inform a statewide effort in NYS that is responsive to community needs and concerns, we conducted a survey of vaccine perceptions in individuals with IDD, their family members (including those who may make decisions on their behalf), and those who work with people with IDD. The aims of the survey were to characterize 1) intent to receive the COVID-19 vaccine; 2) reasons for vaccine hesitancy; and 3) trusted sources of information about the vaccine.

## Methods

The survey was conducted via a cohort design to inform a year-long statewide campaign supported by NYS’s Developmental Disabilities Planning Council (DDPC). Federally funded and mandated Developmental Disability Councils in each US state and territory identify and address the needs of persons with IDD through advocacy and systems change. The DDPC works with the state’s three University Centers of Excellence in Developmental Disabilities (UCEDD), two of which collaborated on this project. This project was deemed exempt from human subjects review by the Office of Human Research Affairs at Einstein College of Medicine-Montefiore Medical Center and the Office for Human Subject Protection at the University of Rochester.

Though comprehensive surveillance data by race and ethnicity for New York’s IDD population are unavailable, data exist for persons receiving Medicaid-funded services through the state’s Office for People with Developmental Disabilities (New York State Department of Health: Office for People with Developmental Disabilities, 2021). The majority have a primary diagnosis of Intellectual Disability (58%), followed by Autism Spectrum Disorder (21%). The most prevalent racial/ethnic groups are White (61%), Black (18%), Hispanic (6%), and Asian (3%).

### Survey Development

The survey was developed by the UCEDD Directors at the two sites (i.e., Bronx and Rochester), who have expertise in survey development, validation, and analysis. To enable comparison with a non-IDD stakeholder community, survey items were either drawn directly or slightly adapted from the Kaiser Family Foundation (KFF) December 2020 Monitoring Survey ^13^. Adaptations included: adding disability-specific concerns to possible reasons for hesitancy; streamlining questions about trusted messengers, and; adding items about supports respondents would need if they chose to get the vaccine. A plain language introduction described the purpose of the survey, and that it was intended for people who: have a disability, spend time with people with disabilities (i.e., family), and/or work with people with disabilities. With a statewide campaign focus on outreach to diverse communities, the survey was translated into Spanish, simplified Chinese, Korean, and Bengali. English, Chinese, Spanish, and Korean survey versions were entered into Research Electronic Data Capture (REDCap), a user-friendly web-based data capture system. Brief blurbs in each language included a link to the survey in REDCap. Data from Bengali respondents were unavailable for analysis.

### Survey Sample

The survey was disseminated via a public link and remained open for response from January 19 - February 9, 2021. The NYS DDPCs and two UCEDDs emailed the survey to their network of partnering organizations, with a request to distribute it to their clients and staff and to post on their website. These organizations included: The InterAgency Council of Developmental Disabilities Agencies, Inc. (a 150+ member organization, including many congregate care providers); local health departments; agencies providing services to children and youth with special health care needs, and Special Olympics New York. Organizations serving persons with IDD who are Hispanic/Latino, Chinese, Korean, and South Asian also distributed the survey. Local and statewide recruitment also occurred through community partner organizations, including school districts, specialty medical clinics, centers on transition for people with IDD, agencies serving people with limited English proficiency, and a statewide Community of Practice on Cultural and Linguistic Competence in IDD.

### Survey Items

A full version of the survey is included in an Appendix.

#### Role

As noted above, respondents could occupy any of three roles with respect to disability: 1) person with IDD; 2) person who spends time with people with IDD (e.g., family); and/or 3) person who works with people with IDD.

#### Vaccine Intent

Intention to receive the COVID-19 vaccine was assessed with the following KFF item: “How likely would you be to get the COVID vaccine if it were offered to you for free?” We combined responses of ‘definitely get,’ ‘probably get,’ and ‘already got the vaccine’ into an ‘Intend/ Got’ category. Responses of ‘probably not get,’ definitely not get,’ and ‘don’t know’ were categorized as ‘Do Not Intend/DK.’ Through branching logic, those who fell within this latter group, alternatively referred to as “Hesitant,” encountered additional questions related to reasons for hesitancy.

#### Reasons for Hesitancy

‘Hesitant’ respondents were asked to rate nine possible reasons for hesitancy as one of the following: a ‘major reason,’ a ‘minor reason,’ ‘not a reason’ or a designation of “unsure/ don’t know.”

#### Decision-Making

Branching logic guided respondents who indicated a role of ‘spend time with people with disabilities’ and who make medical decisions for a person with a disability to report on Vaccine Intent and Reasons for Hesitancy regarding that person with IDD. The items and response options were otherwise identical.

For respondents who provided their zip codes, we used ArcGIS to create maps of where respondents were from, and vaccine preferences by county.

### Analysis

Descriptive statistics were shown as count and percentage for demographics and each item in the survey. Odds ratios and 95% confidence intervals for respondents in the ‘Intend/Got’ group are compared for participants 50+ years old versus 18-49 years old were shown by race. Chi square or Fisher’s exact test was used to test the difference of proportion across roles or race/ethnicity groups. To examine whether the proportion of participants with vaccine preference for self is different from those with vaccine preference for others, McNemar test was used.

### Maps

A 2018 county-level shapefile and tabular datasets containing zip codes with county Federal Information Processing System (FIPS) code was downloaded from the United States Census Bureau and imported into ArcMap. Datasets were joined in ArcMap and the shapefile and tabular data for New York State respondents were exported. Callouts were used to show the distribution of survey respondents in the Northeastern United States and the New York City metropolitan area. To map vaccine preferences, the proportion of “Intend/Got” responses for each of New York’s 62 counties was represented by quintiles on a choropleth map. This map used projections from the NAD 1983 (the state plane coordinate zones for North America) and the FIPS 3102 coordinate system.

## Results

Complete surveys were received from n= 825 individuals, most of whom (85%) spent time with (e.g., family) and/or worked with persons with disabilities (PWD; see Table 1). PWDs represented 11% of the sample. Regarding language, 9% of the sample completed the survey in Spanish, simple Chinese or Korean. The race/ethnicity of the sample is roughly comparable to that of New York state across Asian (9.2% vs. 9% New York State), Black (15.5% vs. 17.6%), Hispanic (15.5% vs. 19.5%), and white (53.8% vs. 69.6%) groups.^25^ Most respondents were women (82.8%).

**Table 1.**
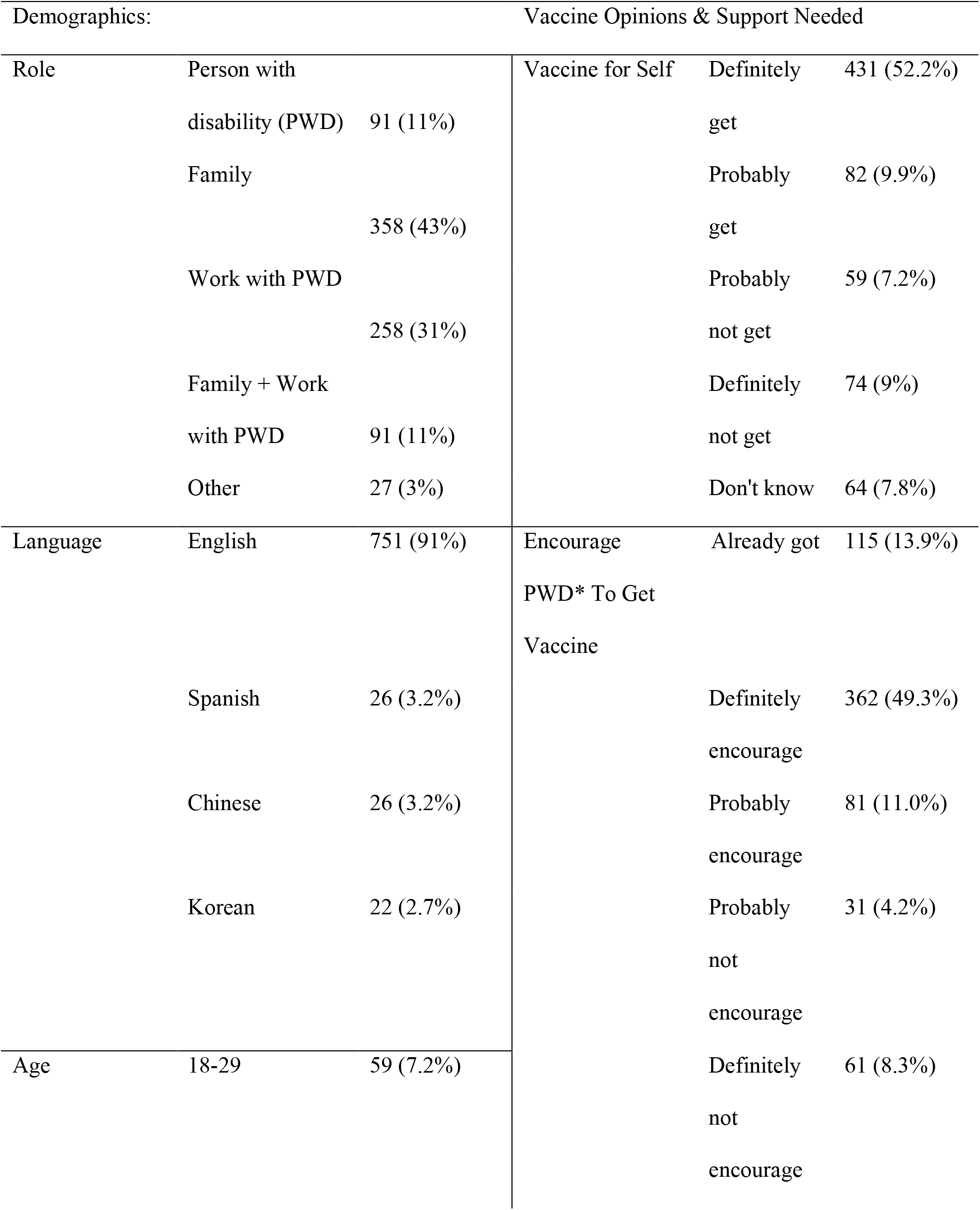
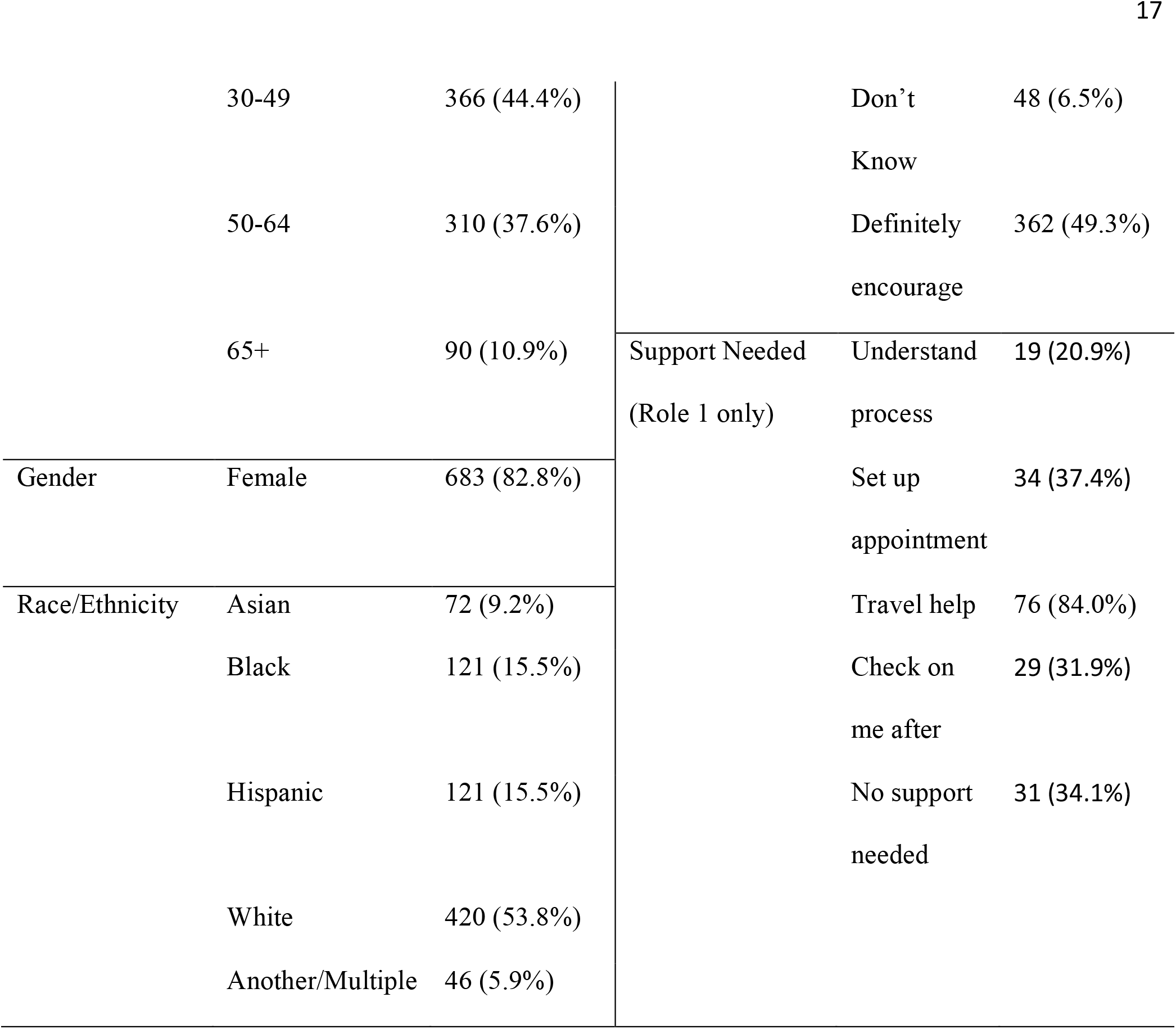
Participant Characteristics and Opinions on Receiving the Vaccine.

In terms of vaccine preference, 62% of respondents would definitely or probably get a COVID-19 vaccine. Further, 13.9% of respondents had already gotten the vaccine-a rate similar to the 13.4% rate for the US adult population at the mid-point of the survey period.^26^ This pattern is similar to that of respondents who make decisions for others. A combined 45% of the sample needed help understanding the process for getting a vaccine and/or setting up the appointment.

In the New York City metropolitan area respondents are clustered in the 5 boroughs of NYC and gradually dispersed throughout Long Island and the NYC suburbs (Figure 1a). Beyond the metro area, most are concentrated in the NYC and Rochester areas in NYS. Several other respondents were observed in Maryland, Washington D.C., and Pennsylvania (Figure 1b). Vaccine preferences are shown in Figure 1c. Responses were received from all 62 of New York State’s counties, though with some counties reporting only a few responses, data should be interpreted with caution. Nevertheless, this map shows higher concentrations of ‘Intend/Got’ responses in the western, central, and southeastern regions of the state.

**Figure 1a-c.**
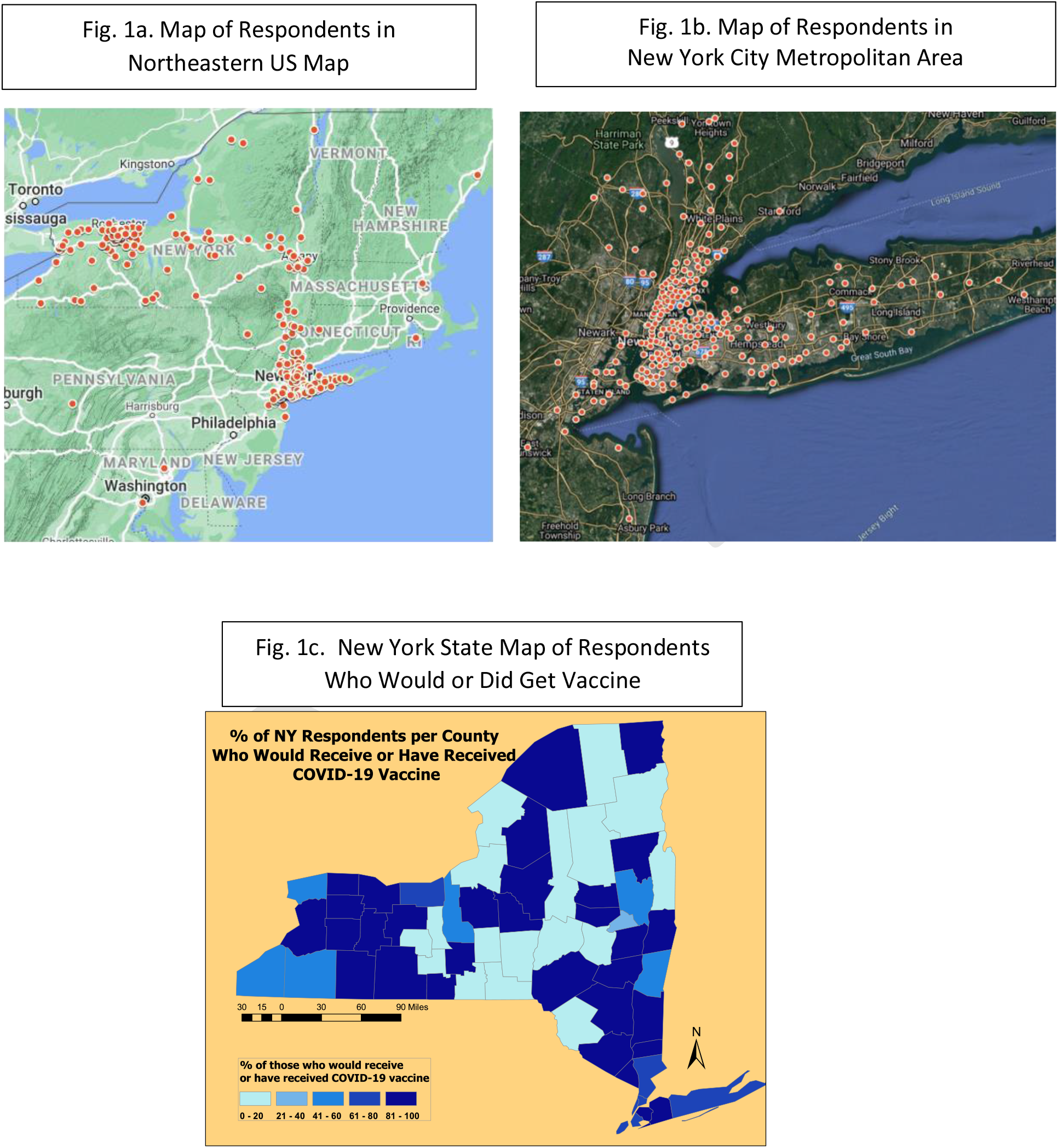
Geographic Maps of Respondents.

The top reasons offered for not getting the vaccine (Figure 2) were side effects (16%), vaccine ‘too new’ (15%), not wanting to be an ‘experiment’ for the vaccine (14%) and not trusting the government (14%). Regarding trusted sources of vaccine information (Figure 3), health professionals ranked highest (92%), followed by friends and family (74%). Newspapers and television were trusted by twice as many people as social media.

**Figure 2.**
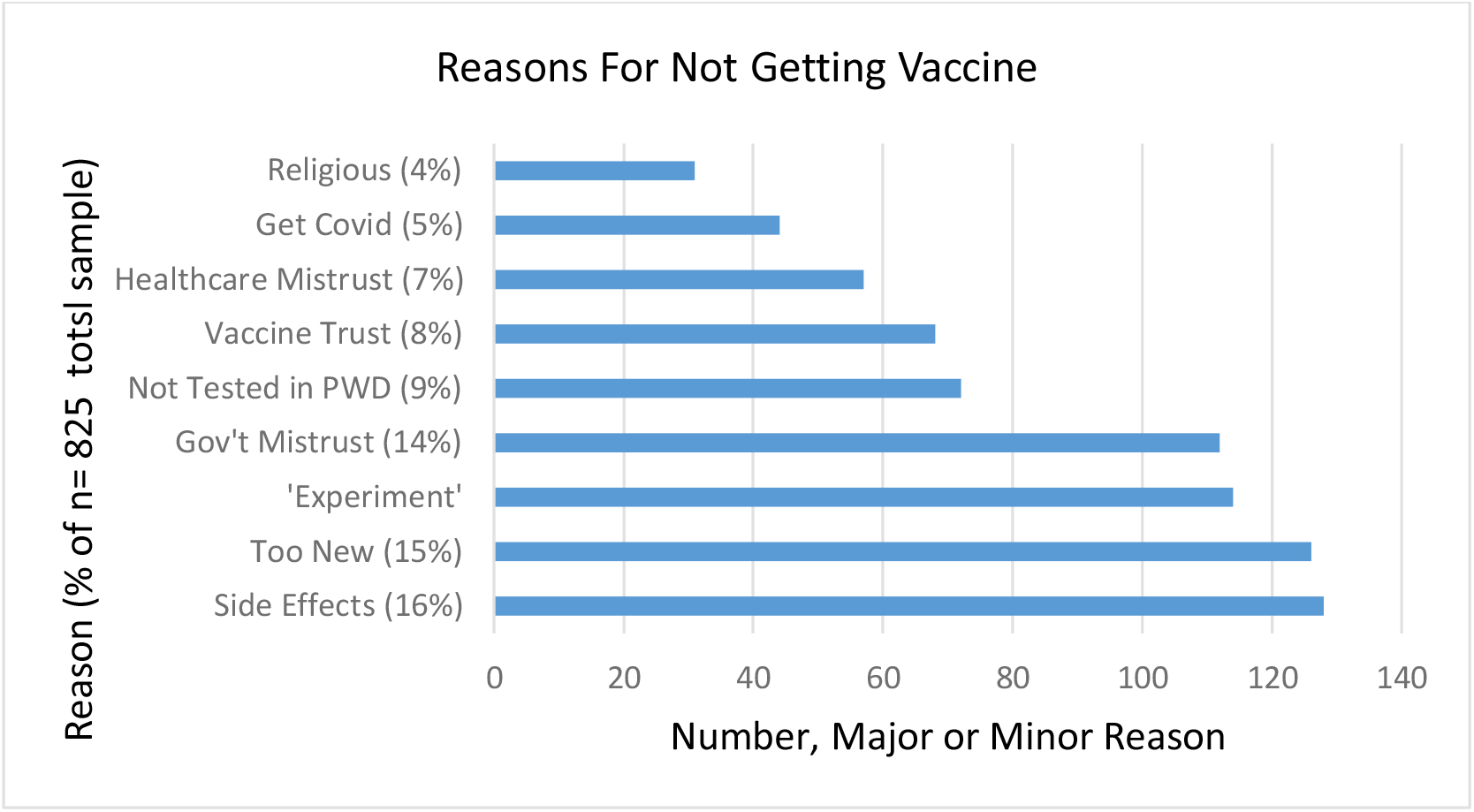
Percentage of Total Sample Respondents Endorsing Reasons to Not Get Vaccine.

**Figure 3.**
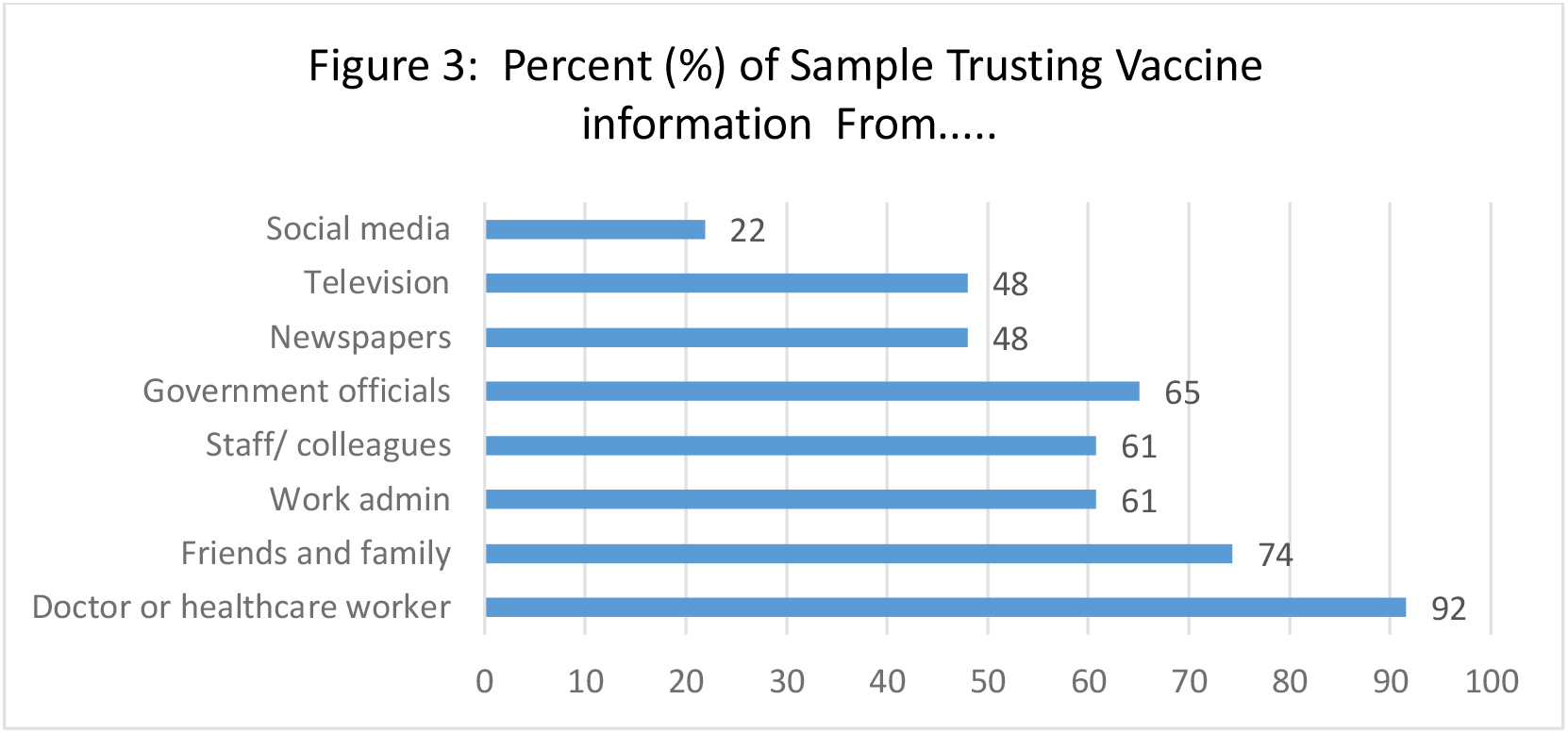
Percentage of Total Sample Endorsing Trusted Messengers of Vaccine Information.

There were no significant differences in report of ‘Intend/Got’ by respondent role (p= 0.28; see Table 2). There were, however, significant differences between preferences for oneself versus for others for whom the respondent makes health decisions (75% vs. 68%; p<.001), as shown in Table 3. Regarding vaccine preferences by race and age (Table 4), Black respondents > 50 years old were more likely to endorse ‘Intend/Got,’ compared to younger respondents (OR= 3.72; 95% CI: 1.73-8.00), as were White respondents (OR= 2.39; 95% CI: 1.36-4.17).

**Table 2:** Preferences for Self, by Role.

| Preferences for Self: |  |  |
| --- | --- | --- |
| Role: | Intend/Got | Do Not Intend/DK |
| Person with Disability<br>(PWD) | 76 (83.5%) | 15 (16.5%) |
| Family Member | 263 (73.5%) | 95 (26.5%) |
| Work with PWDs | 196 (76%) | 62 (24%) |
| Both Family Member &<br>Work with PWD | 73 (80.2%) | 18 (19.8%) |
| Other | 20 (74.1%) | 7 (25.9%) |
| Preference for Others For Whom Respondent is Decision-Maker |  |  |
| Family Member (all) | 443 (60.3%) | 292 (39.7%) |

**Table 3:** Comparison of Vaccine Preference for Self vs. Others (Family Members Only).

|  | Preference for others:<br>Intend/Got | Preference for others:<br>Do Not Intend/DK | Total |
| --- | --- | --- | --- |
| Preference for self:<br>Intend/got | 293 | 38 | 331 (74.9%)* |
| Preference for self:<br>Do Not Intend/ DK | 6 | 105 | 111 (25.1%) |
| Total | 299 (67.7%) | 143 (32.3%) | 442 |

\*p= ≤ 0.0001 (McNemar test).

**Table 4.** ‘Intend/Got’ Vaccine Preference by Race and Age.

| Age | Asian |  | Black |  | Hispanic/ Latino |  | White |  |
| --- | --- | --- | --- | --- | --- | --- | --- | --- |
| in |  |  |  |  |  |  |  |  |
| Years | (freq./ %) | OR<br>(95% CI) | (freq./ %) | OR<br>(95% CI) | (freq./ %) | OR<br>(95% CI) | (freq./ %) | OR<br>(95% CI) |
| 18-49 | 35 (83.3%) | Ref. | 26<br>(37.7%) | Ref. | 61<br>(69.3%) | Ref. | 139<br>(79.4%) | Ref. |
| 50+ | 27 (90.0%) | 1.80<br>(0.43, 7.62) | 36<br>(69.2%) | 3.72<br>(1.73, 8.00) | 25<br>(75.8%) | 1.38 (0.55, 3.46) | 221<br>(90.2%) | 2.39 (1.36, 4.17) |

\* *Note:* Freq. = Frequency; OR = Odds Ratio

Reasons for vaccine concern (major or minor) by race among respondents in the ‘Do Not Intend/DK’ group are shown in Table 5. Concerns about being ‘Used as an Experiment’ differed by race/ethnicity (p<.05) with Hispanic and Black respondents reporting this more often (96% and 91%, respectively) than White or Asian respondents (76% and 67%, respectively). ‘Lack of Trust in Government’ also differed by race (p<.00) with higher rates reported by Black (96%) respondents compared with Hispanic (80%), White (78%), and Asian (0%) respondents. There were no racial/ethnic differences for the remaining seven potential reasons.

**Table 5.** Reasons for Vaccine Concern by Race.

| Major+Minor<br>reason | Asian | Black | Hispanic | White | P-value |
| --- | --- | --- | --- | --- | --- |
| Religious | 1 (33.3%) | 11 (25.0%) | 4 (16.0%) | 8 (21.6%) | 0.72 |
| 'Used as<br>Experiment | 2 (66.7%) | 41 (91.1%) | 24 (96.0%) | 28 (75.7%) | 0.05 |
| Disabilities | 1 (33.3%) | 22 (50.0%) | 18 (72.0%) | 18 (48.7%) | 0.19 |
| COVID | 1 (33.3%) | 19 (42.2%) | 11 (44.0%) | 9 (24.3%) | 0.27 |
| Healthcare Trust | 1 (33.3%) | 21 (47.7%) | 8 (32.0%) | 16 (43.2%) | 0.66 |
| Vaccine Trust | 2 (66.7%) | 22 (50.5%) | 11 (44.0%) | 21 (56.8%) | 0.73 |
| Too New | 2 (66.7%) | 42 (93.3%) | 25 (100%) | 35 (94.6%) | 0.16 |
| Govt | 0 (0.0%) | 43 (95.6%) | 20 (80.0%) | 29 (78.4%) | 0.00 |
| Side Effects | 2 (66.7%) | 44 (97.8%) | 25 (100%) | 36 (97.3%) | 0.09 |

## Discussion

In the context of COVID-related health disparities in New York State, we conducted a statewide survey on COVID vaccine perceptions among people within the IDD community, including individuals with IDD, their family members, and those directly working with people with IDD. Respondents comprised a diverse sample based on race, ethnicity, age, and language. Approximately 25% of respondents reported hesitancy to receive the vaccine themselves, and about 20% indicated hesitancy to encourage a person with IDD to receive the vaccine. These rates are lower than general population surveys conducted during the same time period ^17^. Intention to receive a vaccine did not differ by role, suggesting that materials to increase vaccine confidence could be relevant to all of these groups. In contrast to previous findings, vaccine hesitancy did not differ by race, ethnicity, or primary language. There were notable age-related differences, such that older individuals had more intention to receive the vaccine. There was an interaction effect of age and race, indicating that older Black and White individuals were more likely to have vaccine intention compared to younger Black and White respondents.

The top reasons for hesitancy in our sample--concerns about side effects, concerns that the vaccine was too newly developed, not wanting to feel like an “experiment” for the vaccine, lack of overall trust in the government—are comparable to prior surveys ^17^ In contrast to other surveys, we included an item regarding vaccines not having been tested in PWD and found that nearly one in ten respondents cited this as a concern. While there were no group differences in many of the expressed concerns, Black and Hispanic participants had more concerns about being used as an “experiment,” and Black participants expressed lower trust in the government as compared to other racial/ethnic groups. The addition the “experiment” item was based upon community feedback, as well as previous literature on mistrust of the medical system within communities of color ^27^. Ultimately, feeling like an experiment was one of the primary contributors to vaccine hesitancy, which highlights critical opportunities for addressing this concern through community educational campaigns. It will be important to empathize with concerns, particularly in the context of historical and systemic biases, and then provide evidence-based information to help address misconceptions about the vaccine. Despite the presence of mistrust in medical systems, this may not be an insurmountable barrier ^28^ and utilization of trusted community messengers may increase access to vaccine knowledge.

Effective vaccine confidence campaigns for people with IDD should include addressing logistical barriers, in addition to hesitancy. In exploring needed supports to receive the vaccine, people with IDD indicated that assistance setting up the appointment and understanding the process would be most helpful. Existing materials from NYS related to the COVID-19 vaccine process are lengthy, have a high reading level, and do not include plain language versions; as such, this is a significant opportunity for those engaged in vaccine confidence efforts. Of note, over half of individuals with IDD indicated that they did not require any support.

This project has several strengths. First, survey items were adapted from a previously vetted survey (i.e., from Kaiser Family Foundation), which allowed for comparison with national data. Second, to our knowledge, there is limited data on vaccine preferences and barriers in the IDD community. Third, and related to the prior strength, was the inclusion of disability specific items, including vaccine preferences for respondents who make decisions for a person with a disability. Fourth, was the project’s linguistic diversity, as the survey was translated into four languages and distributed through organizations serving these language users. (The survey was translated into Bengali, but no responses received for this version.) Fifth, the project demonstrates the utility of geospatial analysis for identifying locales to target for vaccine uptake. Finally, the survey distribution process served as a means of engaging a broad, statewide network of community partners, who may be excluded from traditional recruitment efforts.

The project also had several limitations. First, as a convenience sample, findings may not be representative of New York’s disability community by diagnosis or living situation. However, results may reflect biases in who chose to respond. Second, the survey items did not include specifiers for disability category or workplace setting (e.g., residential versus community). Comparisons across these subgroups may have yielded interesting findings, and future work on vaccine perceptions within the disability community should consider including these identifiers. Third, as the survey did not specify a definition of support, it is unclear if respondents who indicated that that did not need support were familiar with actual methods of obtaining an appointment, (e.g., computer registration or telephone access) and then traveling alone to a designated and possibly unfamiliar vaccine site. Fourth, data were analyzed descriptively, rather than through predictive models. The survey was developed primarily to inform messaging for community vaccine confidence campaigns; as such, we did not have a priori hypotheses or specific research questions identified. However, group differences emerged from the dataset suggest that more rigorous evaluation of characteristics that influence vaccine hesitancy would be warranted. Finally, findings were based solely upon self-report of *intention* to receive the COVID-19 vaccine and are not necessarily indicative of actual vaccination behavior. In addition, many of the fears expressed were specific to the COVID-19 vaccine (e.g., rapidity of vaccine development, side effects), which may not be predictive of overall vaccine hesitancy.

## Conclusions

Understanding community perceptions of the COVID-19 vaccine is critical to providing equitable vaccine education that adequately addresses concerns and misconceptions. Statewide data suggest that, in New York, all members of the disability community would benefit from educational materials that address concerns about vaccine development and side effects. Group differences based on age and race/ethnicity indicate the value of tailoring materials and of using trusted community messengers. While the presented data and trends are individual to New York State, the survey process and items can provide a template for similar evaluations in communities for whom vaccine hesitancy is a concern.

## Data Availability

Data will be made available upon request

## Acknowledgments

Thank you to those who assisted with survey translation, including Yali Pang, Miriam Franco, Marcelle Pachter, Rachel Passmore, Claudia Perez, Jackie Hayes, and Leonie Winheller. We also thank our community partners for assistance disseminating the survey.

